# Low-Dose Aspirin Adherence Following Objective cell-free RNA-Based Preeclampsia Risk Testing: A Real-World Survey Study

**DOI:** 10.64898/2026.06.08.26355195

**Authors:** Alison B. Moe, Carrie Haverty, Manfred Lee, Susan E. Hahn, Thomas F. McElrath, Maneesh Jain, Morten Rasmussen, Aimee Corso, Matt L. Larson, Honiko Morrison, Laura M. Melroy, Joseph Roofeh, Barbara Phelps-Sandall, Daniel G. Kiefer, Joseph R. Biggio

## Abstract

**Introduction:** Preeclampsia (PE) is a leading cause of maternal and neonatal morbidity and mortality, and low-dose aspirin (LDA) prophylaxis is the cornerstone of evidence-based prevention. Despite guideline recommendations, LDA adherence remains poor, with 10-25% of moderate-risk patients taking aspirin. Objective personalized risk stratification using biomarkers has been shown to motivate behavior change in other disease contexts. Survey data suggest that patients are more motivated to take aspirin if informed by an objective predictive test. Here, we report real-world LDA adherence among patients who received a high-risk result from a cell-free RNA (cfRNA) PE risk prediction test.

**Methods:** This retrospective, observational survey study included asymptomatic patients of advanced maternal age (AMA; ≥35 years at delivery) with singleton pregnancies without USPSTF-defined preexisting high-risk conditions for PE who received the cfRNA PE risk prediction test. Patients who opted in to receive text message surveys were asked about LDA use following receipt of test results. High adherence was defined as reporting LDA use on at least 6 of 7 days per week at least 85% of the time surveyed. The primary analysis included patients with a high-risk test result and at least one LDA frequency survey response following receipt of test result. The observed proportion of adherent patients was compared to a baseline estimate of 25% using an exact binomial test.

**Results:** Of 166 patients who received a cfRNA PE risk prediction test result, 48 (28.9%) received a high-risk result. Of these, 29 (60%) opted in and responded to at least one survey, constituting the primary analysis population. Twenty-seven of the 29 (93.1%; 95% CI: 78.0– 98.1%) were classified as highly adherent, significantly higher than the 25% baseline adherence estimate for moderate-risk patients (p < 0.0001).

**Conclusion:** Among surveyed patients who received a high-risk cfRNA PE test result, the proportion classified as highly adherent to LDA (93%) substantially exceeded published estimates of adherence in a similar patient population and met the clinically meaningful threshold of ≥80% associated with reduced risk of preterm preeclampsia. These findings indicate that objective and personalized biomarker risk testing may be a powerful driver of behavior change that current guidelines have failed to produce.

**Key Points:**

1. Due to non-specific clinical guidelines, real-world adherence to low-dose aspirin (LDA) prophylaxis in patients at risk of developing preeclampsia is critically low, with 10-25% of moderate-risk patients estimated to take aspirin. This represents a major and persistent gap in preventive care.
2. Among patients who received a high-risk result from the cfRNA PE risk prediction test, 93.1% were highly adherent to LDA use based on self-reported surveys. This exceeds the threshold of ≥80% adherence shown to be associated with a clinically significant reduction in preterm preeclampsia.
3. The observed adherence rate was statistically significantly higher than published estimates of aspirin uptake rates among moderate-risk patients, providing real-world evidence that objective risk assessment based on an individual’s personal biology may substantially improve LDA prophylaxis—a change that guideline-based counseling alone has not been able to achieve.

## 1. Introduction

### 1.1 The Impact of Low-Dose Aspirin Prophylaxis

Preeclampsia (PE) is a hypertensive disorder of pregnancy that affects an estimated 8% of pregnancies in the United States and is a leading cause of maternal and neonatal morbidity and mortality.^1^ Because there is no acceptable cure once PE is established, early risk identification and prophylactic treatment are critical. Low-dose aspirin (LDA) is the most well-established pharmacologic intervention, recommended by the United States Preventive Services Task Force (USPSTF), the American College of Obstetricians and Gynecologists (ACOG), and the Society for Maternal-Fetal Medicine (SMFM).^2-4^ The Aspirin for Evidence-Based Preeclampsia Prevention (ASPRE) trial demonstrated that LDA reduced the incidence of preterm PE in high-risk women by 62%, with preterm PE occurring in only 1.6% of the LDA group compared with 4.3% in the placebo group.^5,6^ This benefit was contingent upon adherence: ASPRE reported that 79.9% of participants took 85% or more of their required tablets, a threshold that can be used as a benchmark for clinically meaningful LDA adherence.^5^

The downstream clinical and economic consequences of adequate LDA adherence are substantial. A secondary analysis of the ASPRE trial data found that adherent LDA use was associated with a 68% reduction in neonatal intensive care unit (NICU) length of stay. This reduction was driven primarily by the prevention of births before 32 weeks of gestation and carries important implications for improved neonatal outcomes and healthcare resource utilization.^6^

### 1.2 Limitations of Current Practice

Despite the strength of the evidence and guideline endorsement, real-world adherence to LDA for PE prevention remains critically poor. A review of the literature shows that LDA is used by fewer than 50% of patients with high-risk factors and fewer than 25% of moderate-risk patients (defined per USPSTF criteria as those with two or more moderate-risk factors for preeclampsia, including nulliparity, advanced maternal age, obesity, Black race, and low socioeconomic status, among others).^3^ This represents a profound quality gap in preventive care. Given that over 80% of pregnant women have at least one moderate-risk factor as defined by the USPSTF,^7^ the inability to identify who among this broad population is truly at elevated risk for preterm PE further complicates targeted counseling and treatment.

It has been demonstrated that USPSTF moderate-risk factors in the absence of high-risk conditions have little value for estimating the risk of developing preeclampsia, leading to nonspecific and inconsistent LDA recommendations, contributing to continuously increasing rates of PE.^8,9^ This further underscores the need for objective, personalized risk stratification tools that are based on an individual’s biology and can meaningfully distinguish high-risk from low-risk individuals within the broad moderate-risk category without dependence on personal characteristics and demographics.

### 1.3 Objective Testing as a Driver of Behavior Change

The impact of biomarker-based tests upon patient behavior has been examined in the context of obstetric care. In a 2024 survey study, 87.9% of respondents reported they would be more motivated to follow their provider’s medication recommendations if they better understood the risks of preeclampsia, with 87.1% of respondents reporting that they would be more motivated to take aspirin specifically if it were recommended following an objective predictive risk test.^10^ However, this data reflected anticipated rather than actual behavior change. A critical open question was whether this stated intent to adhere would translate into real-world behavior following receipt of an objective high-risk test result.

### 1.4 Cell-free RNA (cfRNA) Preeclampsia Risk Prediction Test

The test (Encompass™, Mirvie, San Francisco, CA) is a clinically validated, cell-free RNA (cfRNA) assay used on a blood sample taken between 17.5 and 22 weeks of gestation.^11^ It is designed to identify, months before the onset of symptoms, whether a patient is at high or low risk of developing preterm preeclampsia based on an individualized cfRNA signature.^11^ Preterm preeclampsia is defined as being diagnosed before 37 weeks leading to a preterm birth and/or severe features. Unlike USPSTF-based risk stratification, which relies solely on static clinical and demographic characteristics, the measurement of cfRNA levels reflects the activity of maternal and placental genes, yielding a highly individualized risk prediction. The test has been clinically validated with a sensitivity of 91% and specificity of 74% for predicting preterm preeclampsia in singleton AMA pregnancies without USPSTF-defined preexisting high-risk conditions.^11^

### 1.5 Study Objective

This study was designed to evaluate the real-world decision impact of the cfRNA PE risk prediction test by measuring LDA adherence following receipt of a high-risk result among patients classified as moderate risk per USPSTF criteria. The test’s focus on stratifying pregnant patients with moderate-risk factors is key because this is the population for whom current guideline-based risk stratification provides the least discriminatory value.^8^

Specifically, we sought to determine whether patients’ intention to adhere to LDA prophylaxis following objective preeclampsia risk testing translated into real-world behavior, and whether the observed population-level adherence rate met or exceeded the clinically meaningful threshold of ≥79.9%.^5,6,10^

## 2. Materials and Methods

### 2.1 Study Design

This was an observational retrospective study utilizing de-identified survey responses collected as part of the routine test experience. No experimental procedures, randomization, or blinding were employed. Pursuant to 45 CFR 46.104(d)(4), the Institutional Review Board determined this research project to be exempt from IRB oversight, with a Waiver of Documentation of Informed Consent approved.

### 2.2 Data Source and Survey Administration

The data for this study consisted of de-identified responses to text message surveys sent to patients who opted in to receive such communications at the time of testing. Patients who agreed to receive surveys were sent a series of brief questions via text message at regular intervals following the issuance of their test report. There were no study-specific procedures; all data were collected as part of the standard product experience.

Survey responses were considered eligible for analysis if the response date was at least seven days after the test report was issued. Patients in the primary analysis completed between 1 and 19 post-test LDA adherence surveys, with a mean of 12 completed surveys per patient. Patients were asked whether their provider had recommended aspirin, whether they were currently taking aspirin, and the frequency of aspirin use in the past week. Two sequential versions of the aspirin frequency question were used during the study period. Both versions contained the following introductory language and instructions: “We’re checking in to hear about your aspirin use in the last week. Have you taken it: [response options] Reply with just a letter.” The later version offered the following response options: A) 6–7 days, B) 4–5 days, C) 1–3 days, D) 0 days, E) I’ve stopped aspirin. An earlier version used a slightly different response scale: A) Every day, B) 5–6 days, C) 1–4 days, D) 0 days, E) I’ve stopped aspirin. Adherence classifications were harmonized across both versions as described in Section 2.4.

### 2.3 Study Population

The cfRNA PE risk prediction test is clinically validated for and available to asymptomatic pregnant individuals of advanced maternal age (maternal age ≥35 years at delivery) with a singleton pregnancy and without USPSTF-defined preexisting high-risk conditions (e.g., prior history of preeclampsia, chronic hypertension).^11^ Included patients were seen at a geographically diverse set of obstetric care settings across the United States, including private OB-GYN practices, multispecialty women’s health groups, and academic medical centers. Eligible subjects included patients who: (1) received a high-risk test result; (2) opted in to the text-based survey program; (3) provided at least one post-test survey response regarding aspirin frequency. Patients were excluded if they did not opt in to text message surveys or did not complete any eligible aspirin adherence surveys.

### 2.4 Adherence Definition

Patients were classified as “highly adherent” if they reported near-daily LDA use on at least 85% of their aspirin surveys. This threshold was selected to align with the definition of clinically meaningful adherence from the ASPRE trial, in which “good adherence” was defined as an intake of 85% or more of the required number of tablets.^5^ Applying this threshold longitudinally across multiple survey time points provides a conservative, real-world measure of consistent daily LDA use over the course of the pregnancy.

Due to the two survey versions described in Section 2.2, the adherence threshold was operationalized as follows: for patients who completed the later survey version (n = 15), a response of “6–7 days” was counted as adherent; for patients who completed the earlier survey version (n = 14), responses of “every day” or “5–6 days” were both counted as adherent, reflecting equivalent near-daily use on that scale. A patient was classified as highly adherent if adherent responses comprised at least 85% of their completed surveys.

### 2.5 Statistical Analysis

Descriptive statistics were used to characterize the study population and primary adherence outcome. The observed proportion of highly adherent patients was reported with a 95% confidence interval (CI), calculated using the Wilson score method.

Exact binomial test (one-sided; H□: observed adherence > baseline) was used to assess whether the observed adherence rate significantly exceeded the baseline adherence rate. A p-value < 0.05 was considered statistically significant.

## 3. Results

### 3.1 Study Cohort

Between August 2024 and March 2026, a total of 166 test reports were issued. Of these, 48 patients (28.9%) received a high-risk result, consistent with performance in the test validation cohort.^11^ Among the 48 high-risk patients, 38 (79.2%) opted in to the text-based survey program. Of the 38 enrolled patients, 29 (76.3%) provided at least one post-test aspirin frequency survey response and were included in the primary analysis. The study participant flow is presented in Figure 1.

**Figure 1.**
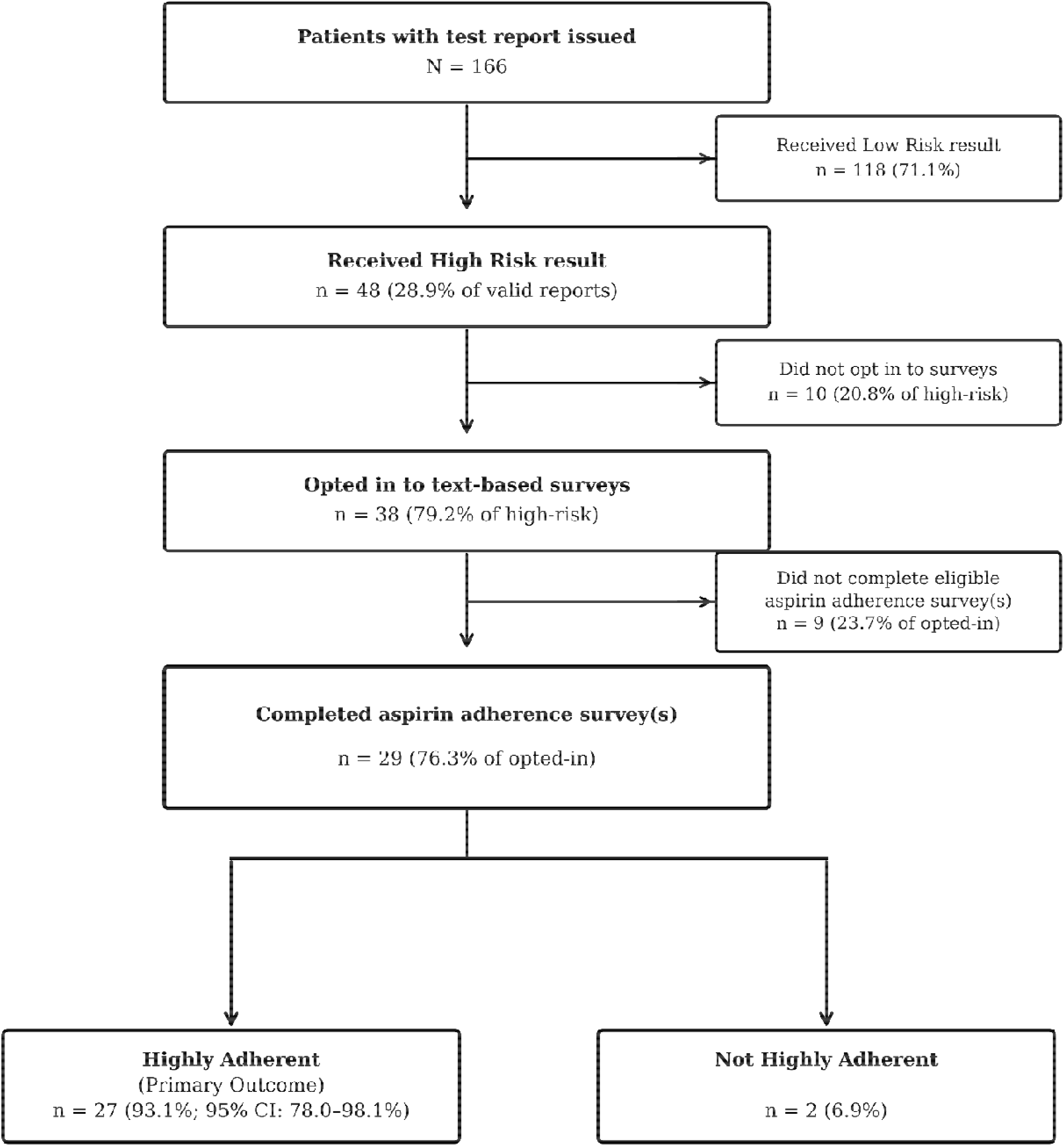
Study CONSORT Diagram. Of 166 patients with a cfRNA preeclampsia risk prediction test report issued, 48 (28.9%) received a high-risk result. Of those, 38 (79.2%) opted in to the text-based survey program. Of enrolled patients, 29 (76.3%) provided at least one post-test aspirin frequency survey response and comprised the primary analysis population. Of these, 27 (93.1%) were classified as highly adherent, defined as reporting aspirin use on at least 6 of 7 days on at least 85% of surveys.

### 3.2 Low-Dose Aspirin Adherence

Of the 29 patients in the primary analysis, 27 (93.1%; 95% CI: 78.0–98.1%) were classified as highly adherent to low-dose aspirin, defined as reporting aspirin use on at least 6 of 7 days at least 85% of the time they were surveyed. This rate meets and exceeds the 79.9% “good adherence” rate observed in the ASPRE trial,^5^ and is substantially higher than published real-world baseline adherence estimates.^3,8^

The remaining 2 patients (6.9%) were not classified as highly adherent under the stringent study definition. One patient reported aspirin use of 4–5 days per week on both eligible adherence surveys, reflecting a stable but slightly below-threshold pattern. The second patient completed 8 qualifying post-test surveys, of which 5 responses indicated aspirin use 6–7 days per week and 3 responses indicated use 4–5 days per week, yielding an overall adherence rate of 62.5% (5/8 surveys). However, both patients reported aspirin intake consistent with moderate adherence by ASPRE trial criteria, defined as intake between 50% and 84.9% of prescribed doses.^5^

### 3.3 Statistical Comparison to Baseline Adherence

The observed adherence rate of 93.1% was significantly higher than the pre-specified baseline comparator of 25% (p < 0.0001), which represents a conservative upper-bound estimate of real-world low-dose aspirin adherence among moderate-risk patients.^3,8,12^

### 3.4 Sensitivity Analyses

As a sensitivity analysis, a stricter definition of highly adherent was applied to the patients who completed the earlier version of the survey, counting only “every day” responses as adherent. Under this stricter definition, 25 of 29 patients (86.2%; 95% CI: 69.4%–94.5%) remained highly adherent, with only 2 patients reclassified as non-adherent.

A second sensitivity analysis addressed potential non-responder bias by conservatively treating all 9 opted-in patients who did not complete any eligible surveys as non-adherent. Under this assumption, 27 of 38 opted-in patients (71.1%; 95% CI: 55.2%–83.0%) were classified as highly adherent, which is still statistically significantly higher than the 25% upper-bound comparator. Extending this assumption further and treating the 10 elevated-risk patients who did not opt in to the survey program as non-highly adherent yields 27 of 48 (56.2%; 95% CI: 42.3%–69.3%) classified as highly adherent. This most conservative possible estimate remains statistically significantly higher (p < 0.0001). Across all sensitivity analyses, the primary conclusion is robus to assumptions about non-responders.

## 4. Discussion

This is a study of patients who received a high-risk cfRNA preeclampsia risk prediction test result as part of their clinical care and therefore represents real-world study conditions. Importantly, receipt of a high-risk result was associated with a high uptake and adherence to LDA prophylaxis at 93.1% (95% CI: 78.0–98.1%). This substantially and significantly exceeds historical baseline adherence estimates in a comparable patient population. The observed rate is also meaningfully higher than the 79.9% of ASPRE trial participants classified as having “good adherence”; this population-level benchmark was associated with a statistically significant reduction in preterm preeclampsia as well as improved neonatal outcomes.^5,6^ Critically, the patients in this study are precisely those for whom current guideline-based risk stratification performs most poorly, namely women who are broadly categorized as “moderate risk” on the basis of AMA, for whom personalized risk assessment offers the greatest value.^8^ These real-world findings confirm what previous patient surveys have found – that objective, personalized, biology-based preeclampsia risk testing may result in a level of patient engagement and treatment adherence that clinical guideline-based counseling alone has failed to achieve.^10,13^

### 4.1 Real-World Adherence in Context

The magnitude of the gap between the observed adherence rate and published population-level comparators warrants emphasis. The Society for Maternal-Fetal Medicine (SMFM) has documented that aspirin is taken by fewer than 25% of patients with multiple moderate-risk factors, even among those who receive a recommendation.^3^ Literature reports that only 24-37% of moderate-risk patients are recommended LDA.^8,12,14^ When combined, these deficits in physician under-prescription and poor adherence mean that fewer than 10% of women who are eligible for and could therefore benefit from LDA prophylaxis actually receive it and take it.^12,14,15^ In contrast to the 25% adherence estimated under optimistic guideline implementation and the 10% observed in real-world practice,^14,15^ the 93.1% adherence achieved in this study approaches the theoretical maximum, differences that are highly statistically significant (p < 0.0001). Importantly, the present study assessed not just adherence but used a conservative definition that required sustained, near-daily use across multiple survey time points during the pregnancy, which is the type of adherence that is clinically meaningful for preeclampsia prevention.^5,6^

The clinical significance of the observed adherence level is further underscored by a recent modeling analysis by Rolnik et al., which used ASPRE and Screening Program for Preeclampsia (SPREE) cohort data to demonstrate that low-dose aspirin’s preventive effect against preterm PE is highly dependent on adherence. The preventive effect diminishes substantially at 50% adherence and becomes statistically non-significant and indistinguishable from placebo below approximately 42%.^16^ The 93.1% adherence observed in this study is well above this threshold, placing it in the range where low-dose aspirin’s full preventive benefit is expected to be realized.

### 4.2 Validation of Prior Survey Findings

The present findings constitute a direct real-world validation of the results previously reported by Cowan et al.^10^ In that prospective survey study, 87.1% of respondents anticipated they would be more motivated to take aspirin if an objective predictive test showed they were at higher risk. The actual adherence rate observed in our study (93.1%) mirrors this anticipated behavior, suggesting that the stated intentions by pregnant (or recently pregnant) people translated into real-world action when patients received a personalized test result.

### 4.3 Parallel Evidence from Other Fields

These results are consistent with a broader body of evidence demonstrating that objective and personalized biomarker-based risk information facilitate informed treatment choices and improve adherence to treatment recommendations across disease contexts. In cardiovascular medicine, coronary artery calcium (CAC) scoring provides a direct quantitative measure of coronary atherosclerosis that has been shown to substantially improve patient motivation for statin therapy and lifestyle modification as compared to when counseled on risk factors alone.^17,18^ In oncology, the Oncotype DX Breast Recurrence Score test has been shown to drive concrete behavior change: patients who receive a low-risk score are significantly more likely to forgo chemotherapy and receive endocrine therapy alone, a treatment shift that traditional clinicopathologic features alone cannot reliably achieve.^19,20^ This concept of individualized, biomarker-based risk testing as a driver of patient engagement is increasingly reflected in clinical guidance; ACOG Clinical Consensus Guideline^21^ explicitly recommends comprehensive individual risk assessment and shared decision-making as foundational to modern prenatal care, signaling a broader paradigm shift away from standardized “one-size-fits-all” approaches to care.

The cfRNA PE risk prediction test occupies an analogous role in obstetrics. For a population of patients who are broadly told they have “moderate risk” based on demographic and clinical characteristics that are individually weakly predictive,^8^ receipt of a high-risk result derived from their own cfRNA profile is a qualitatively different and more actionable form of risk information. This represents a substantial improvement over current guideline-based screening approaches, which have a high test-positive rate but lack individualized risk estimates, limiting their ability to discriminate which patients will go on to develop preeclampsia, particularly preterm preeclampsia. This may explain why the behavior change reported in hypothetical survey data^10^ appears to have materialized into real-world adherence in this study.

### 4.4 Clinical and Health Economic Implications

The clinical importance of closing the aspirin adherence gap extends beyond preeclampsia prevention itself. Wright et al.^6^ demonstrated that aspirin-associated prevention of preterm PE translated into a 68% reduction in NICU length of stay, a difference that carries substantial downstream cost implications. Budget impact analyses of cfRNA-based PE testing have estimated gross savings of approximately $56 million per 10,000 pregnancies tested. This includes approximately $4.6 million in maternal cost savings, with the remainder driven predominantly by avoided NICU costs.^22^ Real-world adherence data of the magnitude reported here are a critical input to such models, as the economic benefit of accurate risk stratification is only realized if high-risk patients actually take their prescribed low-dose aspirin. The 93% adherence observed in this study demonstrates that the clinical utility of cfRNA preeclampsia risk testing extends meaningfully beyond the test result itself, to actual benefits reliant on preventive treatment uptake.

### 4.5 Limitations

These results should be considered within the parameters of the study design. First, the sample size is small (n = 29 in the primary analysis population), which limits the precision of the estimated adherence rate and precludes subgroup analyses. The wide confidence interval reflects the exploratory nature of this study and warrants replication in larger cohorts.

Aspirin adherence was assessed by self-report via text message survey. Self-reported adherence is susceptible to social desirability bias and may overestimate true adherence. While objective verification of adherence (e.g., pill counts) was not used, the use of repeated surveys across multiple time points, with a stringent definition requiring 85% or more of surveys to reflect near-daily use, mitigates some of this concern. Of note, pill counts are themselves subject to the Hawthorne effect; patients who know their medication supply is being physically monitored may alter their behavior as a result of that observation. Survey-based methods are not free of this concern, but the episodic, low-burden nature of text surveys may reduce observational influence compared to daily physical pill tracking.

Additionally, the study population reflects patients who opted in to text-based survey participation, which may introduce selection bias. Patients who opt in to engagement programs may be inherently more motivated to adhere to medical recommendations than those who do not. Notably, however, the opt-in rate (79%) and survey completion rate (60%) are encouraging for a real-world, non-incentivized program, suggesting that the responding population is unlikely to represent a highly self-selected subgroup. Furthermore, the sensitivity analyses that were performed directly address this concern: even under the most conservative assumptions – treating all patients who did not opt in or complete surveys as non-highly adherent — the observed adherence rate remains statistically significantly higher than baseline comparators, suggesting that selection bias alone cannot account for the findings.

The study population was restricted to patients who accessed the cfRNA PE prediction test before 22 weeks gestation and answered surveys conducted in English and may not generalize to all populations, particularly those who did not receive timely prenatal care, non-English speakers, or other underserved communities. Future studies should continue to prioritize inclusion of diverse and underserved populations, particularly given known disparities in preeclampsia outcomes.

This study does not include a concurrent control group of patients who received standard guideline-based counseling without the cfRNA test. The baseline comparators used are external estimates from multiple published sources, but may not perfectly reflect the counterfactual adherence in an otherwise identical population.

## 5. Conclusion

In this real-world survey study, patients who received a high-risk result from the cfRNA preeclampsia risk prediction test demonstrated a low-dose aspirin adherence rate of 93%, validating the adherence data previously reported in survey-based studies.^10^ This is consistent with the clinically meaningful threshold established in the ASPRE trial and statistically significantly higher than estimated population-level comparators in the absence of objective risk testing.^5,8^ While prospective, controlled studies with clinical outcome endpoints are warranted to confirm the downstream impact of this adherence improvement on preeclampsia incidence and neonatal outcomes, these findings provide direct real-world evidence that cfRNA-based PE risk testing results in meaningful patient behavior change. This represents a pivotal shift in prenatal care – moving away from population-level generalized guidelines towards personalized, biomarker-driven risk stratification.

## Data Availability

The data that support the findings of this study are available from the corresponding author upon reasonable request. Inquiries can be directed to.

## References

1. US Preventive Services Task Force. Aspirin Use to Prevent Preeclampsia and Related Morbidity and Mortality: US Preventive Services Task Force Recommendation Statement. JAMA. 2021;326(12):1186–1191. 10.1001/jama.2021.14781.

2. ACOG Committee Opinion No. 743: Low-Dose Aspirin Use During Pregnancy. Obstet Gynecol. 2018;132(1):e44–e52. 10.1097/AOG.0000000000002708.

3. Society for Maternal-Fetal Medicine Patient Safety and Quality Committee, Combs CA, Kumar NR, Morgan JL. Prophylactic low-dose aspirin for preeclampsia prevention – quality metric and opportunities for quality improvement. Am J Obstet Gynecol. 2023;229(2):B2–B9. 10.1016/j.ajog.2023.04.039.

4. Society for Maternal-Fetal Medicine Patient Safety and Quality Committee, Ghartey J, Combs CA. Updated checklists for preeclampsia risk-factor screening to guide recommendations for prophylactic low-dose aspirin: SMFM Special Statement. Pregnancy. 2026;2:e70212. 10.1002/pmf2.70212.

5. Rolnik DL, Wright D, Poon LC, O’Gorman N, Syngelaki A, de Paco Matallana C, Akolekar R, Cicero S, Janga D, Singh M, Molina FS, Persico N, Jani JC, Plasencia W, Papaioannou G, Tenenbaum-Gavish K, Meiri H, Gizurarson S, Maclagan K, Nicolaides KH. Aspirin versus Placebo in Pregnancies at High Risk for Preterm Preeclampsia. N Engl J Med. 2017;377(7):613– 622. 10.1056/NEJMoa1704559.

6. Wright D, Rolnik DL, Syngelaki A, de Paco Matallana C, Machuca M, de Alvarado M, Mastrodima S, Tan MY, Shearing S, Persico N, Jani JC, Plasencia W, Papaioannou G, Molina FS, Poon LC, Nicolaides KH. Aspirin for Evidence-Based Preeclampsia Prevention Trial: Effect of Aspirin on Length of Stay in the Neonatal Intensive Care Unit. Am J Obstet Gynecol. 2018;218(6):612.e1–612.e6. 10.1016/j.ajog.2018.02.014.

7. Wheeler SM, Myers SO, Swamy GK, Myers ER. Estimated Prevalence of Risk Factors for Preeclampsia Among Individuals Giving Birth in the US in 2019. JAMA Netw Open. 2022;5(1):e2142343. 10.1001/jamanetworkopen.2021.42343.

8. McElrath TF, Jeyabalan A, Khodursky A, Moe AB, Lee M, Jain M, Goetzl L, Sutton EF, Simmons PM, Saade GR, Saad A, Pacheco LD, Park-Hwang E, Frolova AI, Carter EB, Collier AY, Kiefer DG, Berghella V, Boelig RC, Elovitz MA, Gyamfi-Bannerman C, Biggio JR, Rood K, Grobman WA, Haverty C, Rasmussen M. Utility of the US Preventive Services Task Force for Preeclampsia Risk Assessment and Aspirin Prophylaxis. JAMA Netw Open. 2025;8(7):e2521792. 10.1001/jamanetworkopen.2025.21792.

9. Fink DA, Kilday D, Cao Z, Larson K, Smith A, Lipkin C, Perigard R, Marshall R, Deirmenjian T, Finke A, Tatum D, Rosenthal N. Trends in maternal mortality and severe maternal morbidity during delivery-related hospitalizations in the United States, 2008 to 2021. JAMA Netw Open. 2023;6(6):e2317641. 10.1001/jamanetworkopen.2023.17641.

10. Cowan A, Haverty C, MacDonald R, Khodursky A. Impact of Early Preeclampsia Prediction on Medication Adherence and Behavior Change: A Survey of Pregnant and Recently-Delivered Individuals. BMC Pregnancy Childbirth. 2024;24(1):196. 10.1186/s12884-024-06397-z.

11. Elovitz MA, Gee EPS, Delaney-Busch N, Moe AB, Reddy M, Khodursky A, La J, Abbas I, Mekaru K, Collins H, Siddiqui F, Nolan R, Boelig RC, Kiefer DG, Simmons PM, Saade GR, Saad A, Carter EB, McElrath TF, Quake SR, DePristo MA, Haverty C, Lee M, Namsaraev E, Berghella V, Collier AY, Frolova AI, Park-Hwang E, Pacheco LD, Sutton EF, Jain M, Rood K, Grobman WA, Biggio JR, Gyamfi-Bannerman C, Jeyabalan A, Rasmussen M. Molecular Subtyping of Hypertensive Disorders of Pregnancy. Nat Commun. 2025;16(1):2948. 10.1038/s41467-025-58157-y.

12. Singh N, Shuman S, Chiofalo J, Cabrera M, Smith A. Missed Opportunities in Aspirin Prescribing for Preeclampsia Prevention. BMC Pregnancy Childbirth. 2023;23(1):717. 10.1186/s12884-023-06039-w.

13. Ahmed S, Brewer A, Tsigas EZ, Rogers C, Chappell L, Hewison J. Women’s attitudes, beliefs and values about tests, and management for hypertensive disorders of pregnancy. BMC Pregnancy Childbirth. 2021;21:665. 10.1186/s12884-021-04144-2.

14. Freret TS, Bryant AS, James KE, Kaimal AJ, Melamed A, Clapp MA. Changes in Low-Dose Aspirin Use After Updated Guidance on Sociodemographic Risk Factors for Preeclampsia. O&G Open. 2025;2(2):e069. 10.1097/og9.0000000000000069.

15. Olson DN, Russell T, Ranzini AC. Assessment of Adherence to Aspirin for Preeclampsia Prophylaxis and Reasons for Nonadherence. Am J Obstet Gynecol MFM. 2022;4(5):100663. 10.1016/j.ajogmf.2022.100663.

16. Rolnik DL, Tan MY, Syngelaki A, Wright D, Poon LC, Nicolaides KH. Combined impact of first-trimester risk and aspirin adherence on preterm preeclampsia prevention. Am J Obstet Gynecol. 2026;234(6):1759–1767. 10.1016/j.ajog.2026.02.014.

17. Rozanski A, Gransar H, Shaw LJ, Kim J, Miranda-Peats L, Wong ND, Rana JS, Orakzai R, Hayes SW, Friedman JD, Thomson LEJ, Polk D, Min J, Budoff MJ, Berman DS. Impact of Coronary Artery Calcium Scanning on Coronary Risk Factors and Downstream Testing: The EISNER Prospective Randomized Trial. J Am Coll Cardiol. 2011;57(15):1622–1632. 10.1016/j.jacc.2011.01.019.

18. Kalia NK, Miller LG, Nasir K, Blumenthal RS, Agrawal N, Budoff MJ. Visualizing Coronary Calcium Is Associated with Improvements in Adherence to Statin Therapy. Atherosclerosis. 2006;185(2):394–399. 10.1016/j.atherosclerosis.2005.06.018.

19. Torres S, Trudeau M, Gandhi S, Warner E, Verma S, Pritchard KI, Petrella T, Hew-Shue M, Chao C, Eisen A. Prospective Evaluation of the Impact of the 21-Gene Recurrence Score Assay on Adjuvant Treatment Decisions for Women with Node-Positive Breast Cancer in Ontario, Canada. Oncologist. 2018;23(7):768–775. 10.1634/theoncologist.2017-0346.

20. Schneider JG, Khalil DN. Why Does Oncotype DX Recurrence Score Reduce Adjuvant Chemotherapy Use? Breast Cancer Res Treat. 2012;134(3):1125–1132. 10.1007/s10549-012-2134-1.

21. American College of Obstetricians and Gynecologists Committee on Clinical Consensus— Obstetrics. Tailored prenatal care delivery for pregnant individuals: ACOG Clinical Consensus No. 8. Obstet Gynecol. 2025;145(5):565–577. 10.1097/AOG.0000000000005889.

22. Hahn S, Lee M, Nathanson BN, Chopra A, Zambelli-Weiner A. RNA-based screening for preterm preeclampsia reduces costs in pregnancies of advanced maternal age. Presented at: SMFM Pregnancy Meeting; 2026; Washington, DC, USA.

